# Effects of front-of-package labels for added sugars and non-sugar sweeteners (NSS) on parents’ perceptions and selections of foods and beverages for their children: a randomized experiment

**DOI:** 10.1101/2025.05.20.25327959

**Authors:** Allison C. Sylvetsky, Mariana F. Grilo, James W. Krieger, Yichen Jin, Sydney Pryor, Lindsey Smith Taillie

## Abstract

**Introduction:** This study aimed to evaluate effects of front-of-package labels (FOPL) for added sugars and non-sugar sweeteners (NSS) on U.S. parents’ selection and perceptions of foods and beverages.

**Methods:** Parents of children 2-12 years old (n= 2,896) completed an online survey and were randomized to: no-FOPL control (n = 990), added sugar FOPL (n = 949), or FOPLs for both added sugars and NSS (n = 957). Participants were shown four fruit drinks and four yogurts. For each set, one product was unsweetened, one was sweetened with added sugars, one with NSS, and one had added sugars and NSS. Products were shown in random order, with FOPLs per assigned treatment arm. Participants also viewed six NSS-containing products separately and reported their purchase intentions and product perceptions.

**Results:** Participants exposed to the added sugar and NSS FOPLs were more likely to choose the unsweetened fruit drink (58% versus 39% for added sugar FOPL (p<0.001) and 32% for control (p<0.001)) and unsweetened yogurt (60% versus 29% for added sugar FOPL arm (p<0.001) and 14% for control (p<0.001)). Participants in the added sugar and NSS FOPL arm were also more likely to identify unsweetened products as the healthiest (70% for fruit drinks, 76% for yogurts), compared to the added sugar FOPL arm (46% for fruit drinks, 49% for yogurts) and control (37% for fruit drinks, 33% for yogurts) (all p<0.001). Those who viewed NSS FOPLs also had lower purchase intentions and perceptions of product healthfulness for NSS products, and higher perceptions of product sweetness compared to the added sugar FOPL or control arms.

**Conclusions:** Viewing products with both added sugar and NSS FOPLs increased the likelihood of choosing the unsweetened option and lowered parents’ purchase intentions and healthfulness perceptions of NSS-containing products. FOPLs for both added sugar and NSS may support parents in making healthier choices for their children.

## INTRODUCTION

Numerous countries worldwide have adopted front-of-package labeling policies for added sugars and other nutrients of public health concern.^1^ In January 2025, the United States (U.S.) Food and Drug Administration (FDA) issued a proposed rule to implement front-of-package labels (FOPLs) for nutrients of public health concern, including added sugar.^2^ A potential unintended consequence is that such a policy may encourage manufacturers to replace added sugars with non-sugar sweeteners (NSS). This has occurred in other countries where only added sugar FOPLs but not NSS labels were required, such as Chile.^3–5^ Increased use of NSS is concerning, because while NSS are considered safe from a toxicological perspective,^6^ accumulating evidence demonstrates that NSS consumption may increase the risk of diet-related chronic disease.^7^

In some countries, such as Colombia, Mexico, and Argentina, the FOPL policy requires a warning label for NSS or a statement disclosing that the product contains NSS. For example, in Mexico, the NNS FOPL reads “Contains sweeteners – not recommended for children.”^8^ A study investigating the impact of the NSS FOPL in Mexico demonstrated that it lowered the perceived healthfulness of an NSS-containing fruit drink,^9^ and a recent analysis of product reformulation following the implementation of the FOPL policy in Mexico demonstrated that its implementation did not lead to increases in NSS use.^10^ Thus, in addition to providing consumers with information that a product contains NSS, NSS FOPLs can also discourage manufacturers from reformulating with NSS.

This is notable because, in 2023, the World Health Organization (WHO) issued a guideline^7^ that NSS should not be used for weight control or to reduce risk of noncommunicable diseases, and there is continued uncertainty and accumulating concern about health effects of NSS, particularly among children. Even prior to the WHO guideline, several public health organizations had put forth recommendations indicating that NSS should not be consumed by children.^11^ Despite these recommendations, however, the use of NSS is increasing in the global food supply^12^ and NSS are widely found in products consumed by children.^13,14^ Consumers, including parents of young children, have difficulty recognizing NSS in food and beverage products^14,15^ and often do not realize that they and/or their children consume NSS.^15^ For example, a study that assessed parents’ perceptions of NSS in beverages found that less than one-third of parents who provided NSS-containing fruit drinks or flavored waters to their child accurately identified the product as containing NSS.^16^

Despite these increasing concerns about NSS consumption as well as the increasing inclusion of NSS in FOPL policies, few studies have examined whether an NSS FOPL impacts consumer behaviors. To our knowledge, only one prior study has examined how a NSS FOPL influences U.S. parents’ perceptions of NSS-containing products, which was conducted among parents of young children one-to-five years old.^17^ This prior study demonstrated that a FOPL disclosing the presence of NSS, added sugar, and percent juice content increased parents’ ability to identify products with NSS and reduced their perceptions of products’ healthfulness.^17^ However, how the NSS FOPL influenced parents’ product selection was not investigated and the label format and wording tested did not reflect FOPLs currently implemented globally or proposed by the FDA. Another recent study, conducted among adults in Australia, demonstrated that a NSS FOPL, along with a sugar FOPL, avoided an increase in selection of NSS-containing beverages, ^18^ but it is unclear if these results are replicable among US parents.

Lastly, an important question relates to how consumers will perceive and select products with a FOPL system that includes both added sugar and NSS. One concern is that the presence of an NSS label could potentially confuse consumers^8^ and increase selections of products with added sugar, which would detract from efforts to lower added sugar intake. An important question is the degree to which parents will make tradeoffs about different types of sweeteners (sugar, NSS) when making product selections for their children. To have maximum health impact and support an overall healthy dietary pattern, a FOPL system would decrease selection of both NSS-containing products and products with added sugar, but how viewing both added sugar FOPLs and NNS FOPLs influences consumer perceptions and behaviors is currently unknown.

The purpose of this online randomized experiment was to examine how viewing both NSS FOPLs and added sugar FOPLs impact parents’ food and beverage product perceptions and selections for their child, compared with FOPL for added sugars only or no FOPLs (control). We also examined how viewing both added sugar FOPLs and NSS FOPLs influence parents’ purchase intentions and their perceptions of the healthfulness and sweetness of NSS-containing food and beverage products.

## METHODS

### Participants

Parents of children 2-12 years old were recruited from throughout the United States (U.S.) to participate in an online experiment, which took place in November 2024. Using data from a similar online experiment comparing effects of FOP natural claims on parents’ purchase intentions,^19^ we estimated that 698 participants per arm (n=2,094 total) were required to provide 80% power to detect differences in the odds of selecting the unsweetened product between the three treatment arms.

Recruitment was carried out using CloudResearch Prime Panels, a national survey panel. Inclusion criteria were having at least one child between 2-12 years old, residing in the U.S., being at least 18 years of age and able to read and speak English, and reporting being the primary decision maker for their household’s food and beverage purchases. Eligible individuals were invited to participate and provided with a link to provide informed consent electronically and then complete the online survey administered using Qualtrics™. Recruitment quotas for sex, race, and ethnicity were used to enroll a diverse sample of parents consistent with national census data. The study protocol was reviewed and approved by the Institutional Review Board (IRB) at the George Washington University and all participants provided informed consent prior to beginning the study procedures.

### Stimuli

The designs of the FOPLs (**Supplemental Figure 1**) were informed by a FOPL style tested by the U.S. FDA for added sugar,^20^ and focus group discussions about NSS FOPLs conducted with U.S. parents (manuscript currently under review). The FOPL for added sugar read “high in added sugar” and the FOPL for NSS read “contains non-sugar sweeteners.” The term “non-sugar sweeteners” was used because it encompasses both “artificial” (e.g., aspartame, sucralose) and “natural” (e.g., stevia, monk fruit extract) NSS^17^ and is consistent with the terminology used in recent WHO guidance.^7^ In the control arm, a bar code was used as a neutral label to control for the effect of having a label and to ensure that the branding and packaging were similarly obscured across all treatment arms.

In the choice experiment (see **Procedures** below), participants were shown two sets (fruit drinks and yogurts) of four products: one that was sweetened with NSS and did not contain added sugar, one that contained added sugar and did not contain NSS, one that contained both added sugar and NSS, and one that was unsweetened (contained neither added sugar nor NSS). These two product categories were selected because fruit drinks and yogurts are widely consumed by children,^21,22^ are commonly perceived as healthy by parents,^23,24^ and are available both with and without added sugar and NSS. Images of the fruit drink and yogurt products are available in the **Supplemental Figure 2.**

In the single product assessments (see **Procedures** below), participants viewed six additional NSS-containing products (fruit drink, sweetened milk, flavored water, yogurt, protein bar, canned beans) one at a time. The six products were chosen because they represented different types of products that often contain NSS and included several foods and several beverages.

Three of the products were sweetened with only NSS and three contained both NSS and added sugars **(Supplemental Figure 3)**.

All products shown were branded products that are available outside of the U.S. This approach was selected to reflect products that consumers encounter in real life, increasing generalizability, while also reducing the likelihood that parents would have pre-existing preferences for certain brands.

### Procedures

After providing informed consent electronically, parents were randomized in Qualtrics™ to one of three study arms: 1) a control arm with no FOPLs, 2) an added sugar FOPL or 3) an added sugar and NSS FOPL. Participants first completed the choice experiment, during which, they viewed a set of four fruit drink products and a set of four yogurt products. For each product set, all participants viewed the same four products, shown in random order, but the products had different FOPLs based on participants’ assigned treatment arm. For each set, participants were instructed to select which of the four products they would buy for their children and asked which was the healthiest, most unhealthy, and sweetest. Participants then completed single product assessments, which allowed us to examine how the FOPLs impacted participants’ perceptions of NSS-containing products. Participants reported their purchase intentions and perceptions of products’ healthfulness for their children and perceptions of product sweetness using a 5-point Likert scale (strongly agree, somewhat agree, neither agree nor disagree, somewhat disagree, strongly disagree).

Participants were also asked to answer demographic questions and report whether they and their child habitually consume products with NSS and/or products within the product categories shown in the experiment. Upon completion of the survey, participants received compensation in the amount and form (e.g., gift cards, points) they agreed upon with CloudResearch Prime Panels.

### Measures

The study outcomes and hypotheses were pre-registered on AsPredicted.Com (#198117) prior to enrolling participants and beginning data collection. The primary study outcome was the selection of the unsweetened product for their child during the choice experiment (binary outcome variable). The secondary outcomes included selection of an NSS-sweetened product for their child and selection of a product with added sugar for their child, during the choice experiment (binary outcome variables), as well as intentions to purchase NSS-sweetened products and perceived healthfulness of NSS-sweetened products for children, from the single product assessments (measured on a Likert scale). The perceived sweetness of NSS-sweetened products, during the single product assessment, was examined as exploratory outcomes, and was also measured on a Likert scale. The wording of the questions and response options are available in **Supplemental Table 1.**

We hypothesized that, compared to the control and added sugar FOPL arms, participants who viewed products with FOPLs for both NSS and added sugars would be more likely to select the unsweetened product for their child and less likely to select products with NSS for their child.

### Statistical Analyses

Descriptive statistics, including means ± standard deviation and frequencies (percentage), were used to describe participants’ characteristics and survey responses. For the choice experiment, logistic regression was used to assess differences in the likelihood of choosing the unsweetened, NSS-sweetened, or added sugar-sweetened product(s) between the treatment arms, and the predicted probabilities in each arm were calculated. For the primary outcome (selection of the unsweetened product in the choice experiment), we also tested for interactions by education, race, and sex. Differences in parents’ perceptions of product healthfulness and sweetness were examined using the Chi-square test. For the individual product assessments, ANOVA was used to examine the differences in mean Likert scale ratings across treatment arms, and linear regression was performed to compare Likert scale ratings between the NSS FOPL arm and the control arm, as well as between the two FOPL arms (NSS FOPL versus added sugar only FOPL). We also examined the influence of the FOPLs across different product types to explore the hypothesis that the labels might have differential effects on purchase intentions, perceived healthfulness, and perceived sweetness depending on the type of product, and also explored interactions across education, race, and sex. All analyses were performed using R version 4.3.3, and a p-value < 0.05 was considered to be statistically significant.

## RESULTS

A total of 2,896 parents of children 2-12 years old completed the online randomized experiment. A participant flowchart is available in the **Supplemental Figure 4** and characteristics of the sample are shown, by treatment arm, in **Table 1**. The characteristics of the participants were similar across the three treatment arms; participants were, on average, 40.7 years old, and 53% were female, 78% were Non-Hispanic White, and 15% were Hispanic. Approximately 36% of the participants had received a college degree, and 64% reported habitually consuming foods and/or beverages with NSS at least once per week. Nearly half (46%) reported that their child habitually consumed foods and/or beverages with NSS at least once per week.

**Table 1.** Sociodemographic characteristics of the sample, overall and by treatment arm, n=2,896.

|  | Control<br>n=990 | Added sugar<br>FOPL<br>n=949 | NSS and added<br>sugar FOPL<br>n=957 |
| --- | --- | --- | --- |
| Sex, n (%) |  |  |  |
| Female | 546 (55.2) | 501 (52.8) | 494 (51.6) |
| Male | 437 (44.1) | 444 (46.8) | 460 (48.1) |
| Non-binary | 4 (0.4) | 3 (0.3) | 3 (0.3) |
| Prefer not to answer | 3 (0.3) | 1 (0.1) | 0 (0.0) |
| Age, mean ± SD n=2881 | 40.2 ± 11.3 | 41.3 ± 11.5 | 40.6 ± 11.0 |
| Education (%) |  |  |  |
| Elementary or middle | 29 (2.9) | 32 (3.4) | 21 (2.2) |
| High school graduate | 302 (30.5) | 292 (30.8) | 299 (31.2) |
| Some college | 299 (30.2) | 311 (32.8) | 275 (28.7) |
| College graduate | 240 (24.2) | 211 (22.2) | 256 (26.8) |
| Advanced degree | 120 (12.1) | 103 (10.9) | 106 (11.1) |
| Ethnicity (%) |  |  |  |
| Hispanic | 154 (15.6) | 150 (15.8) | 140 (14.6) |
| Non-Hispanic | 834 (84.2) | 797 (84.0) | 817 (85.4) |
| Prefer not to answer | 2 (0.2) | 2 (0.2) | 0 (0.0) |
| Race (%) |  |  |  |
| White | 775 (78.3) | 736 (77.6) | 733 (76.6) |
| Black | 115 (11.6) | 128 (13.5) | 124 (13.0) |
| Asian | 28 (2.8) | 24 (2.5) | 46 (4.8) |
| Pacific Islander | 2 (0.2) | 1 (0.1) | 0 (0.0) |
| Native American | 8 (0.8) | 10 (1.1) | 9 (0.9) |
| Multiracial | 45 (4.5) | 38 (4.0) | 37 (3.9) |
| Other | 11 (1.1) | 8 (0.8) | 7 (0.7) |
| Prefer not to answer | 6 (0.6) | 4 (0.4) | 1 (0.1) |
| Child's age, mean ± SD | 8.4 ± 3.0 | 8.4 ± 3.1 | 8.4 ± 3.0 |

### Choice experiment

A greater percentage of participants who viewed fruit drinks with both added sugar and NSS FOPLs selected the unsweetened option (58.0%), compared to those seeing only an added sugar FOPL (39.4%) or the control (31.6%) (both comparisons p<0.001) (**Figure 1 and Supplemental Table 2**). For yogurt, 60.5% of participants in the added sugar and NSS FOPL arm selected the unsweetened option, compared to 29.0% in the added sugar FOPL only arm and 14.3% in the control arm (both comparisons p<0.001).

**Figure 1.**
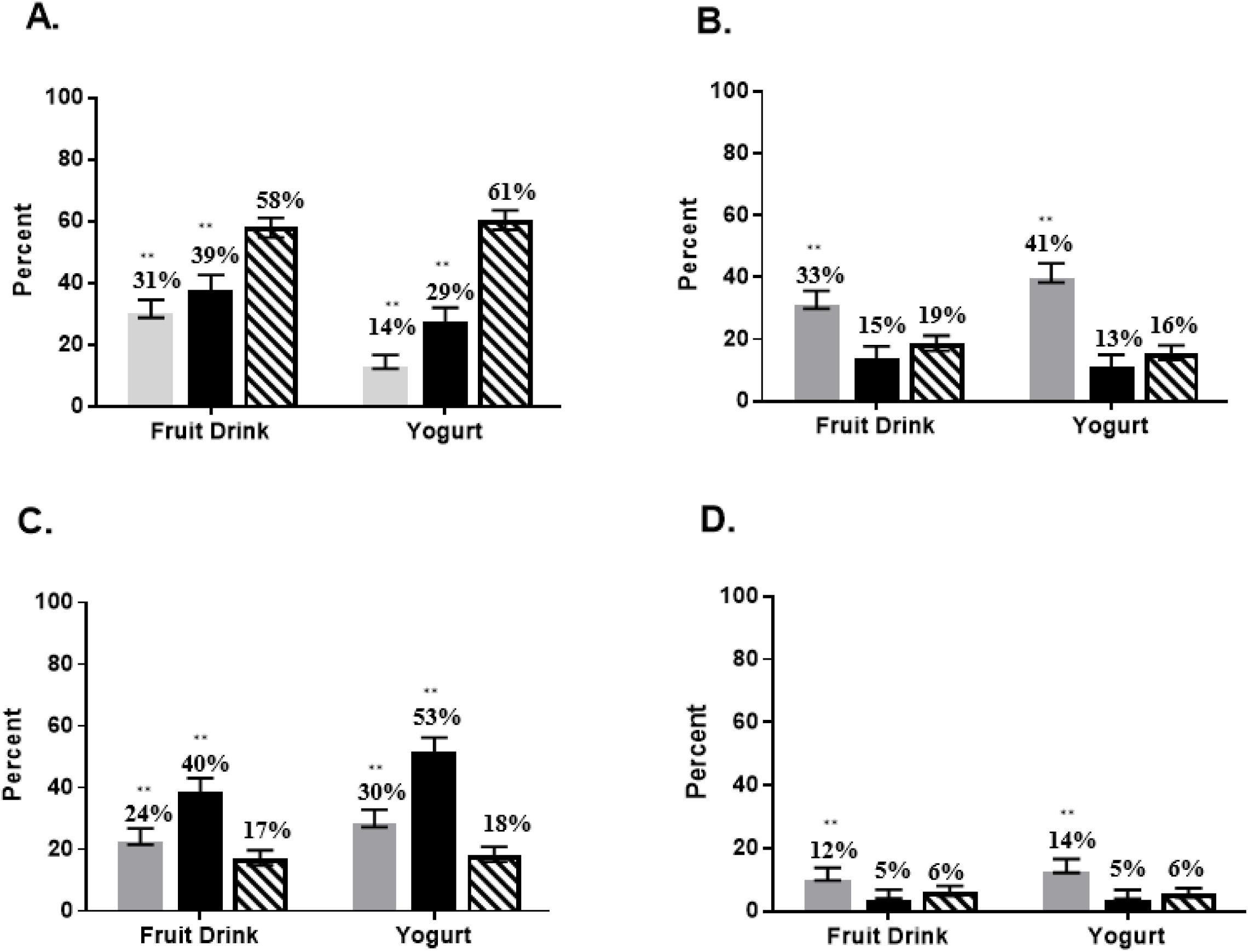
Predicted probability of choosing the unsweetened product (A), product with added sugar (B), product with non-sugar sweeteners (C), or product with both added sugar and non-sugar sweeteners (D), by label arm (n=2,896) Legend: Predicted probabilities (with 95% confidence intervals) of participants selecting the unsweetened product (A, top left), product with added sugar (B, top right), product with non-sugar sweeteners (C, bottom left), or product with both added sugar and non-sugar sweeteners (D, bottom right) for their child in an online randomized choice experiment (n = 2,896). Solid gray bar: no-FOPL control arm. Solid black bar: added sugar FOPL only arm. Gray patterned bar: NSS FOPL + added sugar FOPL arm. NSS: Non-sugar sweeteners. Bars reflect 95% confidence intervals. **p<0.001 compared to added sugar + NSS label group.

There was an interaction between education and the likelihood of selecting the unsweetened fruit drink (p = 0.04); those with a college degree showed weaker associations between labeling arm and purchasing the unsweetened fruit drink, compared to the other education categories (**Supplemental Table 3**). No interactions were observed by race or sex for fruit drinks, and for yogurt, there were no statistically significant interactions (p for interaction > 0.05 for all).

Participants who viewed products with the added sugar FOPL only were more likely to choose products sweetened with NSS (but without added sugar) for their child (**Supplemental Table 2**), for both fruit drinks (40% in the added sugar FOPL arm versus 17% in the NSS FOPL and added sugar FOPL arm (p<0.001) and 24% in the control (p<0.001)) and yogurts (53% in the added sugar FOPL arm versus 18% in the NSS FOPL and added sugar FOPL arm (p<0.001) and 30% in the control, (p<0.001)). However, viewing the NSS FOPL did not result in a higher likelihood of choosing the product with added sugar compared with the added sugar FOPL only arm (18.6% in the NSS FOPL and added sugar FOPL arm versus 15.4% for added sugar FOPL only for fruit drinks, p = 0.062; 15.6% in the NSS FOPL and added sugar FOPL arm versus 12.8% for added sugar FOPL only for yogurts, p = 0.078).

Among participants in the both added sugar and NSS FOPL arm, 70.2% identified the unsweetened product as the healthiest for fruit drinks, compared to 45.6% in the added sugar FOPL arm and 37.5% in the control arm (p<0.001) (**Table 2**). Similarly, for yogurts, 76.3% of participants in the added sugar and NSS FOPL arm identified the unsweetened yogurt as the healthiest option, compared to 49.3% in the added sugar FOPL arm and 32.6% in the control arm (p<0.001). Participants in the added sugar FOPL arm were more likely to identify the product sweetened with NSS (and not added sugar) as the healthiest for both fruit drinks (40.5% in the added sugar FOPL arm, compared with 13.7% in the added sugar and NSS FOPL arm and 25.3% in the control, p<0.001) and yogurts (40.7% in the added sugar FOPL arm, compared with 12.3% in the added sugar and NSS FOPL arm and 29.1% in the control, p<0.001).

**Table 2.**
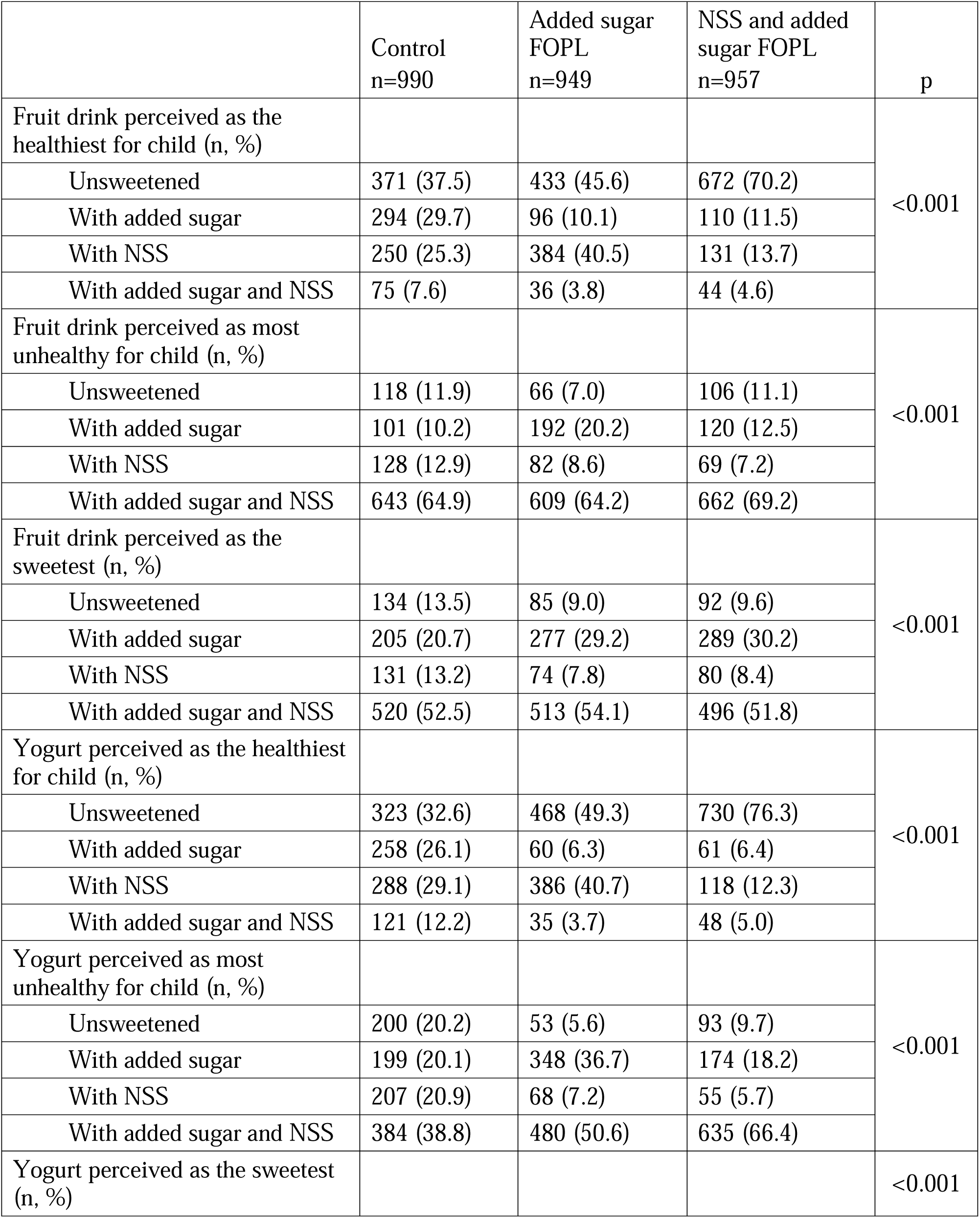
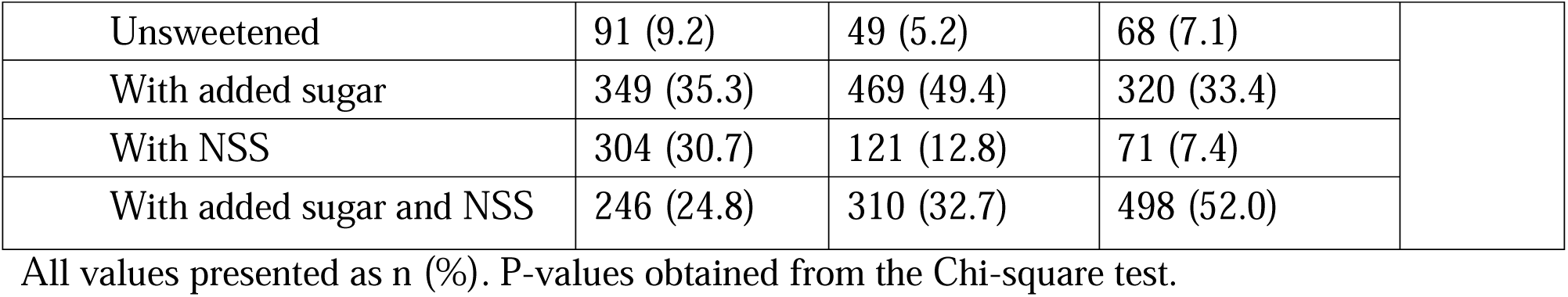
Parents’ perceptions of product healthfulness and sweetness, by treatment arm, n=2,896.

Participants who viewed the products with added sugar and NSS FOPLs were also more likely to select products with both added sugar and NSS as the unhealthiest option, particularly for yogurts (66.4% in the added sugar and NSS FOPL arm versus 50.6% for added sugar only and 38.8% for control, p<0.001). Participants who viewed products with added sugar and NSS FOPLs were also more likely to select the yogurt containing both added sugar and NSS as being the sweetest (52.0% in the added sugar and NSS FOPL arm versus 32.7% for added sugar only and 24.8% for the control, p<0.001), though this pattern was not observed for fruit drinks.

### Single product assessments

Participants who viewed NSS-containing products, that did not contain added sugar, with NSS FOPLs had lower purchase intentions, lower perceptions of product healthfulness, and higher agreement that the product was too sweet for their children, compared to those randomized to the added sugar FOPL or control arm (**Table 3).** For example, when shown yogurt sweetened with only NSS, participants who viewed the product with the NSS FOPL indicated lower purchase intentions (3.10 ± 1.48 for participants who viewed the NSS FOPL versus 4.12 ± 1.02 in the added sugar FOPL arm and 3.98 ± 1.07 in the control, p<0.001), lower perceived healthfulness (3.02 ± 1.43 for participants who viewed the NSS FOPL versus 4.15 ± 0.92 in the added sugar FOPL arm and 4.04 ± 0.95 in the control, p<0.001), and higher agreement that the product was too sweet for their children (3.22 ± 1.30 for participants who viewed the NSS FOPL versus 2.56 ± 1.29 in the added sugar FOPL arm and 2.75 ± 1.29 in the control, p<0.001), compared to those in the added sugar FOPL arm or control. Findings were similar for the flavored water and beans.

**Table 3.** Parents’ intentions to purchase NSS-containing products and their perceptions of product healthfulness and sweetness for their children, measured using 5-point Likert scale, by treatment arm, n=2,896.

|  | Control<br>n=990 | Added sugar<br>FOPL<br>n=949 | NSS and added<br>sugar FOPL<br>n=957 | p |
| --- | --- | --- | --- | --- |
| **NSS-containing products without added sugar** |  |  |  |  |
| **Flavored water** |  |  |  |  |
| Purchase intentions | 3.69 ± 1.15 | 3.73 ± 1.21 | 2.87 ± 1.45 | <0.001 |
| Healthfulness for child | 3.81 ± 1.05 | 3.78 ± 1.11 | 2.71 ± 1.45 | <0.001 |
| Too sweet for child | 2.74 ± 1.28 | 2.66 ± 1.30 | 3.29 ± 1.33 | <0.001 |
| **Yogurt** |  |  |  |  |
| Purchase intentions | 3.98 ± 1.07 | 4.12 ± 1.02 | 3.10 ± 1.48 | <0.001 |
| Healthfulness for child | 4.04 ± 0.95 | 4.15 ± 0.92 | 3.02 ± 1.43 | <0.001 |
| Too sweet for child | 2.75 ± 1.29 | 2.56 ± 1.29 | 3.22 ± 1.30 | <0.001 |
| **Beans** |  |  |  |  |
| Purchase intentions | 3.12 ± 1.42 | 3.34 ± 1.37 | 2.78 ± 1.49 | <0.001 |
| Healthfulness for child | 3.32 ± 1.25 | 3.48 ± 1.17 | 2.80 ± 1.40 | <0.001 |
| Too sweet for child | 2.50 ± 1.26 | 2.49 ± 1.27 | 2.94 ± 1.37 | <0.001 |
| **NSS-containing products with added sugar** |  |  |  |  |
| **Protein bar** |  |  |  |  |
| Purchase intentions | 3.27 ± 1.32 | 2.50 ± 1.38 | 2.39 ± 1.45 | <0.001 |
| Healthfulness for child | 3.37 ± 1.23 | 2.41 ± 1.35 | 2.32 ± 1.37 | <0.001 |
| Too sweet for child | 3.10 ± 1.21 | 3.71 ± 1.26 | 3.66 ± 1.29 | <0.001 |
| **Fruit drink** |  |  |  |  |
| Purchase intentions | 3.19 ± 1.32 | 2.54 ± 1.38 | 2.33 ± 1.42 | <0.001 |
| Healthfulness for child | 2.87 ± 1.30 | 2.22 ± 1.30 | 2.15 ± 1.33 | <0.001 |
| Too sweet for child | 3.57 ± 1.17 | 3.98 ± 1.17 | 3.86 ± 1.26 | <0.001 |
| **Sweetened milk** |  |  |  |  |
| Purchase intentions | 3.35 ± 1.30 | 2.83 ± 1.37 | 2.46 ± 1.41 | <0.001 |
| Healthfulness for child | 3.24 ± 1.25 | 2.61 ± 1.37 | 2.34 ± 1.38 | <0.001 |
| Too sweet for child | 3.15 ± 1.23 | 3.58 ± 1.28 | 3.70 ± 1.29 | <0.001 |

All values presented as mean ± SD. P-values obtained from the ANOVA test.
Likert scale response options: strongly agree (5), agree (4), neither agree nor disagree (3), somewhat disagree (2), strongly disagree (1)

For products with both NSS and added sugars, participants who viewed both FOPLs reported lower purchase intentions, lower perceptions of product healthfulness, and higher agreement that the product was too sweet for their children, compared with the control (**Table 4**). For example, when shown a sweetened milk that had NSS and added sugar, participants who viewed both FOPLs reported lower purchase intentions (2.46 ± 1.41 for participants in the combined FOPL arm and 2.83 ± 1.37 in the added sugar FOPL arm versus 3.35 ± 1.30 in the control, p<0.001), lower perceived healthfulness (2.34 ± 1.38 for participants in the combined FOPL arm and 2.61 ± 1.37 in the added sugar FOPL arm versus 3.24 ± 1.25 in the control, p<0.001), and higher agreement that the product was too sweet for their children (3.70 ± 1.29 for participants in the combined FOPL arm and 3.58 ± 1.28 in the added sugar FOPL arm versus 3.15 ± 1.23 in the control, p<0.001), compared to the control. Findings were similar for the fruit drink and protein bar.

The effect of the NSS FOPL on purchase intentions, perceived healthfulness, or perceived sweetness of the products did not vary by any of the moderators examined (i.e., race, sex of child, education, all moderation p > 0.05).

## DISCUSSION

The findings of this randomized online experiment demonstrate that parents are the most likely to select unsweetened products when viewing both added sugar and NNS FOPLs, compared to no FOPLs or only added sugar FOPLs. Viewing the NNS FOPL and added sugar FOPL, in combination, also lowered parents’ purchase intentions for NSS-containing products and reduced their perceptions of the products’ healthfulness. The magnitude and consistency of the effects of viewing the combination of added sugar FOPLs and NSS FOPLs across the products was striking, and may be explained by the NSS FOPL providing parents with novel, and perhaps unexpected, information about the presence of NSS in the products.^25^

Prior research has demonstrated that parents are often unable to accurately identify products with NSS, and report that they do not want to provide their child with products that contain them.^15^ While FOPLs influence consumer decision making through several different channels,^25^ one pathway is through cognitive effects such as providing knowledge and changing consumer perceptions of products. In a prior study that tested effects of a FOPL for added sugar, NSS, and juice content with parents of young children one to five years old,^17^ viewing the FOPL increased parents’ accuracy in identifying the presence or absence of NSS and lowered parents’ perceived healthfulness of NSS-containing beverages. Similarly, viewing the NSS FOPL implemented in Mexico lowered the perceived healthfulness of a fruit drink among both children and adults.^9^

Consistent with these prior findings,^9,17^ parents who viewed products with NSS FOPLs in the present study reported lower perceived healthfulness of NSS-containing products, regardless of whether the product was sweetened with only NSS or contained both NSS and added sugar.

These results are policy-relevant because the NSS FOPL tested in this online experiment used wording analogous to that used in the NSS FOPL required in Mexico, which reads “contains NSS, not recommended for children.” This is notable because reductions in products containing NSS, along with reductions in nutrients of public health concern, were recently reported following implementation of the FOPL policy in Mexico.^10^ Therefore, in addition to the present findings showing that NSS FOPLs influence consumers and encourage them to choose healthier (i.e., unsweetened) products, NSS FOPLs also discourage manufacturers from reformulating products to contain NSS, thereby reducing their presence in the food supply.

Importantly, viewing a NSS FOPL did not result in a higher likelihood of choosing products with added sugar; rather, viewing a NSS FOPL, in addition to the added sugar FOPL, encouraged selection of healthier (i.e., unsweetened) products. In contrast, those who viewed only the added sugar FOPL, and not the NSS FOPL, were much more likely to perceive the NSS-containing product as the healthiest and select the NSS-containing option. This is consistent with the real-world example of what has occurred in Chile, where increases in NNS were reported following implementation of a policy that required added sugar FOPLs, but did also include NSS FOPLs.^4,5^ While it has been suggested that FOPLs for both added sugar and NSS may confuse consumers,^8^ the present findings demonstrate that FOPLs for both added sugar and NSS help consumers to identify healthier products and promote healthier product choices. These findings are also in contrast to a recent study conducted among university students in Australia, in which, incorporation of a NSS FOPL was shown to reduce the impact of a sugar FOPL unless additional front-of-package nutrition information (i.e., the voluntary Australian health star rating) was also provided.^18^

It is also notable that the effect of the NSS FOPL on parents’ selection of the unsweetened option was more pronounced for yogurts compared with fruit drinks. This may be explained by expectancy disconfirmation theory, which posits that new and unexpected information impacts consumers attitudes towards a product.^25^ While parents may already realize that fruit drinks contain added sugar and/or NSS, the presence of added sugar and/or NSS in yogurts may be more surprising, and thus, the label may have a stronger effect.

Limitations of the study include testing a small selection of products and use of a survey panel, which may limit generalizability. While numerous FOPL formats are used globally, our experiment only tested a single added sugar FOPL and a single NSS FOPL design, and other FOPL wording and format should be examined in future experiments. Furthermore, the lack of a real-world food shopping context (e.g., product placement, pricing information, advertising) and the brief exposure to added sugar and NSS FOPLs in an online experiment may not reflect the influence of FOPLs on product perceptions and selections in real-life.

The strengths of this study include the large and diverse sample of parents and the randomized controlled trial design. Testing added sugar FOPLs and NSS FOPLs with wording consistent with that already used in other countries also enhances the relevance of the study findings. In addition, the use of a neutral control (barcode) label in the control arm allowed us to account for the effect of having a label, and the use of real branded products that are unavailable in the U.S. enhanced generalizability while also reducing confounding by pre-existing brand preferences. Furthermore, we tested both food and beverage products and included foods and beverages sweetened with only NSS and that contained both NSS and added sugar in the single product assessments.

## CONCLUSIONS

The present findings demonstrate that the addition of a NSS FOPL encourages parents to choose unsweetened food and beverage products for their children and does not increase their likelihood of selecting products high in added sugar. Additional research examining effects of different NSS FOPL label formats and investigating NNS FOPL in a real-world food retail setting are needed to inform FOPL policies in the U.S. and globally.

## Data Availability

All data produced in the present study are available upon reasonable request to the authors.

## ACKNOWLEDGEMENTS

This study was funded by a grant from Healthy Eating Research (PI: Sylvetsky). Allison C. Sylvetsky has engaged in consulting work for Spindrift Beverage Co. Inc. and is also engaged in consulting work on behalf of Abbott. The other authors do not have any conflicts of interest to disclose. ACS, JK, and LST conceptualized the project. MFG and SP curated the data. YJ conducted the data analysis. ACS wrote the first draft of the manuscript. All authors reviewed and edited the manuscript.

