## Supplemental Table 1 for "Effects of front-of-package labels for added sugars and non-sugar sweeteners (NSS) on parents’ perceptions and selections of foods and beverages for their children: a randomized experiment"

**Supplemental Table 1.** Wording of questions and response options for choice experiment and single product assessments.

| **Item** | **Response scale** |
| --- | --- |
| **Fruit Drinks: Choice Experiment** |  |
| **Now, you will see four different fruit drinks and will be asked to answer a few questions.**  **When the question refers to your child, please respond for your oldest child who is between 2-12 years old.**  *Order of images presented was randomized.* |  |
| If you had to choose one, which product would you purchase for your child? | 1 = (image of unsweetened fruit drink)  2 = (image of fruit drink with added sugar)  3 = (image of fruit drink with NSS)  4 = (image of fruit drink with added sugar and NSS) |
| In your opinion, which one of these products is the healthiest? | 1 = (image of unsweetened fruit drink)  2 = (image of fruit drink with added sugar)  3 = (image of fruit drink with NSS)  4 = (image of fruit drink with added sugar and NSS) |
| In your opinion, which one of these products is the most unhealthy? | 1 = (image of unsweetened fruit drink)  2 = (image of fruit drink with added sugar)  3 = (image of fruit drink with NSS)  4 = (image of fruit drink with added sugar and NSS) |
| Which of these products is the sweetest? | 1 = (image of unsweetened fruit drink)  2 = (image of fruit drink with added sugar)  3 = (image of fruit drink with NSS)  4 = (image of fruit drink with added sugar and NSS) |
| **Yogurts: Choice Experiment** |  |
| **Now, you will see four different yogurts and will be asked to answer a few questions.**  **When the question refers to your child, please respond for your oldest child who is between 2-12 years old.**  *Order of images presented was randomized.* |  |
| If you had to choose one, which product would you purchase for your child? | 1 = (image of unsweetened yogurt)  2 = (image of yogurt with added sugar)  3 = (image of yogurt with NSS)  4 = (image of yogurt with added sugar and NSS) |
| In your opinion, which one of these products is the healthiest? | 1 = (image of unsweetened yogurt)  2 = (image of yogurt with added sugar)  3 = (image of yogurt with NSS)  4 = (image of yogurt with added sugar and NSS) |
| In your opinion, which one of these products is the most unhealthy? | 1 = (image of unsweetened yogurt)  2 = (image of yogurt with added sugar)  3 = (image of yogurt with NSS)  4 = (image of yogurt with added sugar and NSS) |
| Which of these products is the sweetest? | 1 = (image of unsweetened yogurt)  2 = (image of yogurt with added sugar)  3 = (image of yogurt with NSS)  4 = (image of yogurt with added sugar and NSS) |
| **Single Product Assessment** |  |
| **Now, you will see several different products and be asked to answer questions about each one.**  **When the question refers to your child, please respond for your oldest child who is between 2-12 years old.**  *Order of the products was randomized.* |  |
| “I would purchase this product for my child next week if it was available.” | 1 = Strongly agree  2 = Somewhat agree  3 = Neither agree nor disagree  4 = Somewhat disagree  5 = Strongly disagree |
| “This product is healthy for my child to consume every day.” | 1 = Strongly agree  2 = Somewhat agree  3 = Neither agree nor disagree  4 = Somewhat disagree  5 = Strongly disagree |
| “This product is too sweet for my child to consume every day.” | 1 = Strongly agree  2 = Somewhat agree  3 = Neither agree nor disagree  4 = Somewhat disagree  5 = Strongly disagree |

NSS = Non-sugar sweetener
