## Supplemental Table 2 for "Effects of front-of-package labels for added sugars and non-sugar sweeteners (NSS) on parents’ perceptions and selections of foods and beverages for their children: a randomized experiment"

**Supplemental Table 2**. Predicted probabilities of selecting fruit drink and yogurt products for their children, by treatment arm, n=2,896

|  | Control | Added Sugar FOPL | Added Sugar + NSS FOPL |
| --- | --- | --- | --- |
| **Fruit drinks** | | | |
| Unsweetened | 31.6% (28.8, 34.6)^**^ | 39.4% (36.4, 42.6)^**^ | 58.0% (54.8, 61.1) |
| Contain added sugar | 32.7% (29.9, 35.7)^**^ | 15.4% (13.2, 17.8) | 18.6% (16.3, 21.2) |
| Contain NSS | 24.0% (21.5, 26.8)^**^ | 40.0% (36.9, 43.1)^**^ | 17.1% (14.9, 19.7) |
| Contain added sugar + NSS | 11.6% (9.8, 13.8)^**^ | 5.3% (4.0, 6.9) | 6.3% (4.9, 8.0) |
| **Yogurt** | | | |
| Unsweetened | 14.3% (12.3, 16.7)^**^ | 29.0% (26.2, 32.0)^**^ | 60.5% (57.3, 63.5) |
| Added sugar | 41.4% (38.4, 44.5)^**^ | 12.8% (10.8, 15.0) | 15.6% (13.4,18.0) |
| NSS | 30.0% (27.2, 32.9)^**^ | 53.1% (50.0, 56.3)^**^ | 18.3 (16.0, 20.9) |
| Added sugar + NSS | 14.2% (12.2, 16.6)^**^ | 5.2% (3.9, 6.8) | 5.6% (4.4, 7.3) |

NSS: Non-sugar sweetener. All values reflect predicted probability (95% confidence interval), calculated from logistic regression models examining difference in the likelihood of selecting fruit drink or yogurt products between the treatment arms.

^*^p<0.01 compared to the added sugar + NSS FOPL group.

^**^p<0.001 compared to the added sugar + NSS FOPL group.
