## Supplemental Table 3 for "Effects of front-of-package labels for added sugars and non-sugar sweeteners (NSS) on parents’ perceptions and selections of foods and beverages for their children: a randomized experiment"

**Supplemental Table 3**. Predicted probabilities of selecting fruit drink and yogurt products for their children by treatment arm, stratified by educational attainment

|  | Control | Added Sugar FOPL | Added Sugar + NSS FOPL |
| --- | --- | --- | --- |
| **Fruit drinks: High school or below, n=975** | | | |
| Unsweetened | 25.4% (21.0, 30.3)^***^ | 34.0% (29.0, 39.2)^***^ | 53.4% (48.0, 58.8) |
| Added sugar | 36.0% (31.0, 41.3)^***^ | 18.2% (14.4, 22.8) | 21.9% (17.7, 26.7) |
| NSS | 25.1% (20.7, 30.0) | 41.1% (35.8, 46.5)^***^ | 19.1% (15.1, 23.7) |
| Added sugar + NSS | 13.6% (10.3, 17.7)^***^ | 6.8% (4.5, 10.1) | 5.6% (3.6, 8.8) |
| **Fruit drinks: Some college, n=885** | | | |
| Unsweetened | 32.4% (27.4, 38.0)^***^ | 44.1% (38.6, 49.6)^***^ | 62.6% (56.7, 68.1) |
| Added sugar | 34.1% (29.0, 40.0)^***^ | 12.2% (9.0, 16.4) | 12.7% (9.3, 17.2) |
| NSS | 21.4% (17.1, 26.4) | 40.8% (35.5, 46.4)^***^ | 16.7% (12.8, 21.6) |
| Added sugar + NSS | 12.0% (8.8, 16.2) | 2.9% (1.5, 5.5)^**^ | 8.0% (5.3, 11.9) |
| **Fruit drinks: College or higher, n=1036** | | | |
| Unsweetened | 36.7% (31.8, 41.8)^***^ | 40.4% (35.2, 46.0)^***^ | 58.6% (53.4, 63.5) |
| Added sugar | 28.6% (24.2, 33.5)^**^ | 15.6% (12.0, 20.1) | 20.2% (16.4, 24.6) |
| NSS | 25.3% (21.1, 30.0)^**^ | 37.9% (32.7, 43.4)^***^ | 15.8% (12.4, 19.9) |
| Added sugar + NSS | 9.4% (6.8, 12.9)^*^ | 6.1% (3.9, 9.3) | 5.5% (3.6, 8.4) |
| **Yogurt: High school or below, n=975** | | | |
| Unsweetened | 11.1% (8.2, 15.1)^***^ | 25.6% (21.1, 30.7)^***^ | 58.4% (53.0, 63.7) |
| Added sugar | 42.3% (37.1, 47.7)^***^ | 13.0% (9.7, 17.1) | 14.7% (11.2, 19.0) |
| NSS | 30.0% (25.2, 35.1)^**^ | 55.9% (50.4, 61.2)^***^ | 20.6% (16.5, 25.4) |
| Added sugar + NSS | 16.6% (13.0, 21.0)^***^ | 5.6% (3.5, 8.6) | 6.3% (4.1, 9.5) |
| **Yogurt: Some college, n=885** | | | |
| Unsweetened | 13.7% (10.3, 18.1)^***^ | 29.6% (24.8, 34.9)^***^ | 62.6% (56.7, 68.1) |
| Added sugar | 47.8% (42.2, 53.5)^***^ | 11.6% (8.5, 15.6) | 16.4% (12.5, 21.1) |
| NSS | 28.8% (23.9, 34.2) | 54.7% (49.1, 60.1)^***^ | 17.5% (13.4, 22.4) |
| Added sugar + NSS | 9.7% (6.8, 13.6)^**^ | 4.2% (2.4, 7.1) | 3.6% (2.0, 6.6) |
| **Yogurt: College or higher, n=1036** | | | |
| Unsweetened | 17.8% (14.2, 22.1)^***^ | 31.9% (26.9, 37.2)^***^ | 60.8% (55.7, 65.7) |
| Added sugar | 35.3% (30.5, 40.4)^***^ | 13.7% (10.3, 18.0) | 15.8% (12.4, 19.9) |
| NSS | 31.1% (26.5, 36.1)^***^ | 48.7% (43.2, 54.3)^***^ | 16.9% (13.3, 21.2) |
| Added sugar + NSS | 15.8% (12.4, 20.0)^***^ | 5.7% (3.6, 8.9) | 6.6% (4.5, 9.7) |

^*^p<0.05 compared to the added sugar + NSS FOPL group.

^**^p<0.01 compared to the added sugar + NSS FOPL group.

^***^p<0.001 compared to the added sugar + NSS FOPL group.

NSS: Non-sugar sweeteners. All values reflect predicted probability (95% confidence interval), calculated from logistic regression models examining difference in the likelihood of selecting fruit drink or yogurt products between the treatment groups.
