## Supplemental Figure 1 for "Effects of front-of-package labels for added sugars and non-sugar sweeteners (NSS) on parents’ perceptions and selections of foods and beverages for their children: a randomized experiment"

**Supplemental Figure 1.** Added sugar FOPL, non-sugar sweetener (NSS) FOPL, and control labels shown on products in online randomized experiment.

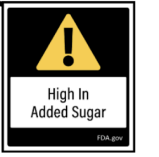

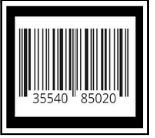

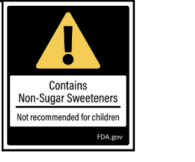
