## Supplemental Figure 2 for "Effects of front-of-package labels for added sugars and non-sugar sweeteners (NSS) on parents’ perceptions and selections of foods and beverages for their children: a randomized experiment"

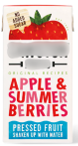

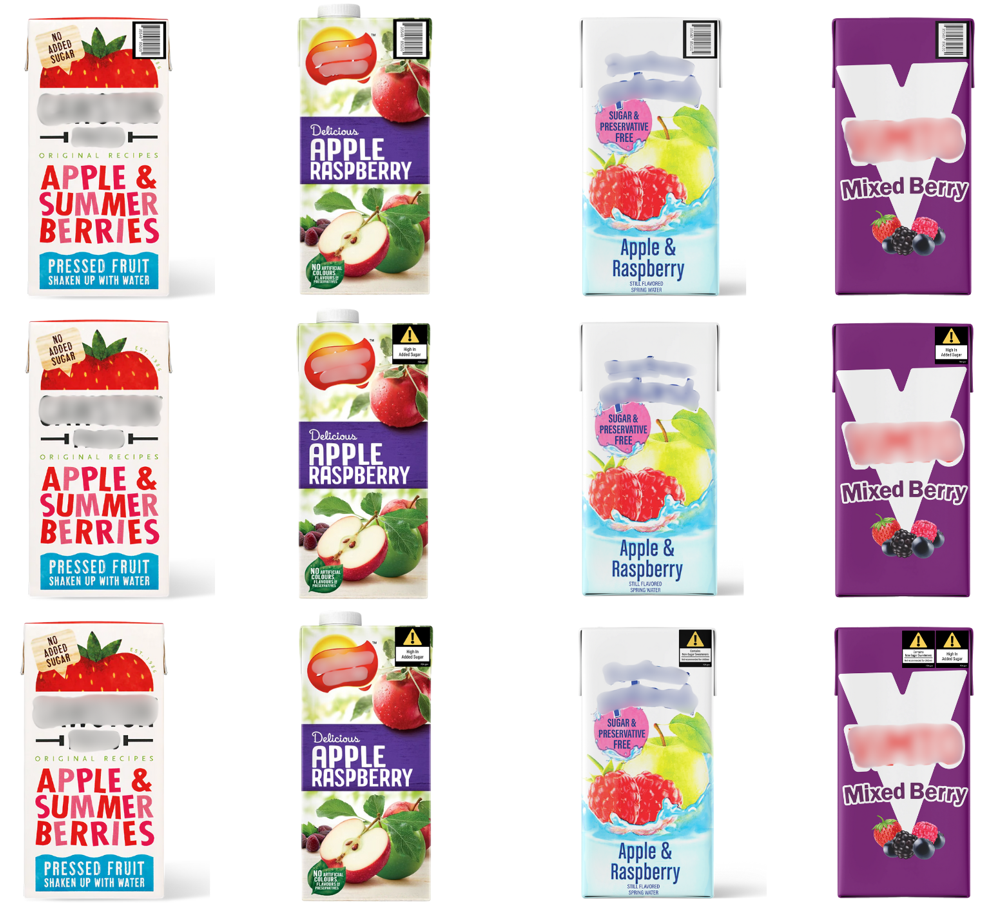

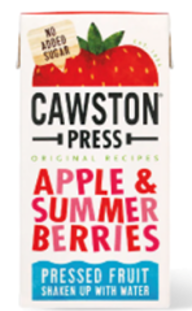

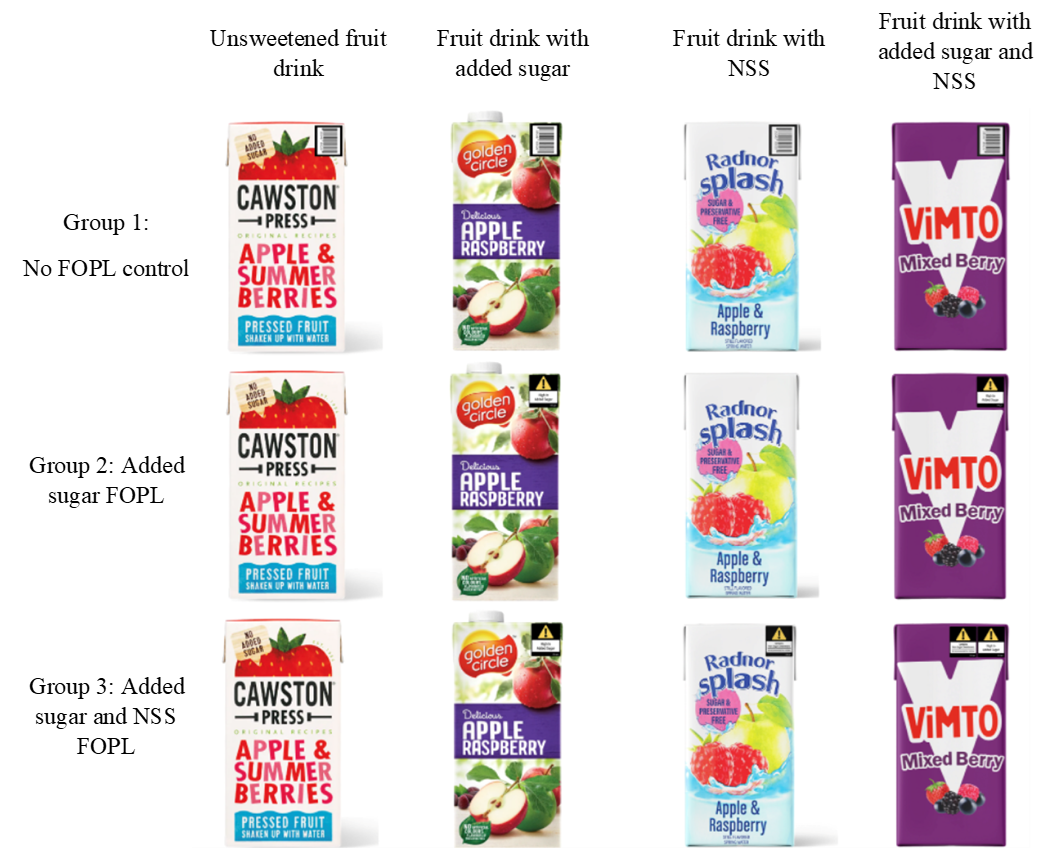
**Supplemental Figure 2.** Fruit drink and yogurt products shown in the online randomized experiment, by treatment arm

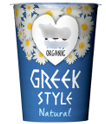

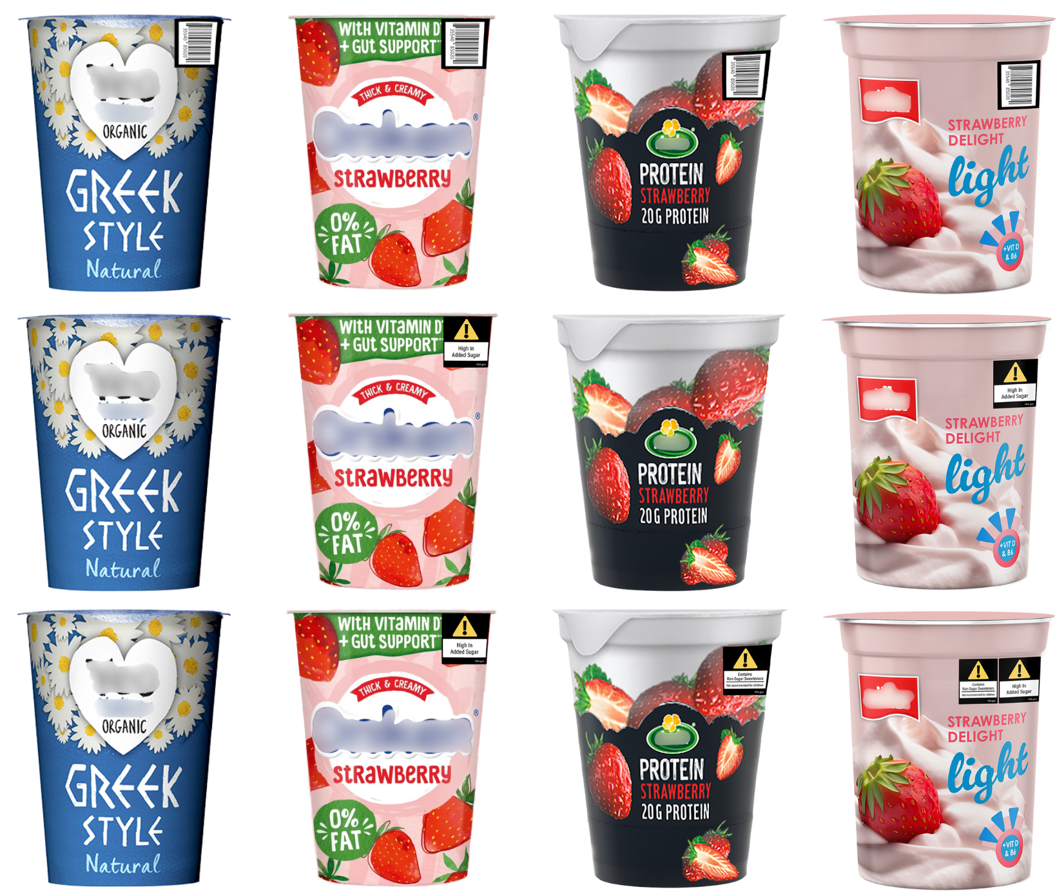

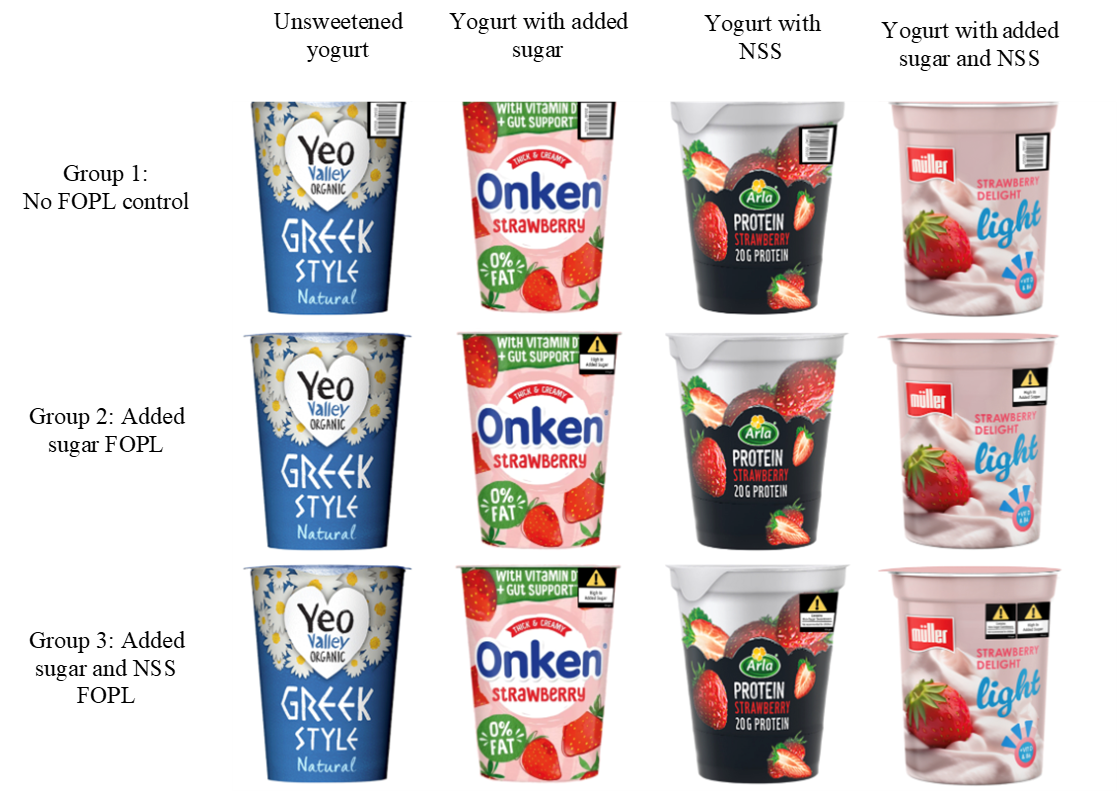
