## Supplemental Figure 3 for "Effects of front-of-package labels for added sugars and non-sugar sweeteners (NSS) on parents’ perceptions and selections of foods and beverages for their children: a randomized experiment"

**Supplemental Figure 3.** Products containing non-sugar sweeteners shown in single product assessments

| **Control Group** | **Added Sugar FOPL** | **NSS FOPL and Added Sugar FOPL** |
| --- | --- | --- |
| Contains NSS and not added sugar | | |
| 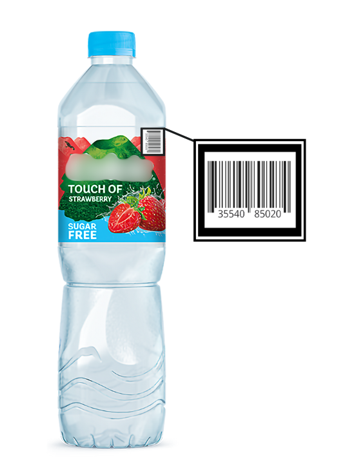 | 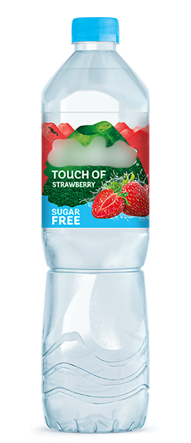 | 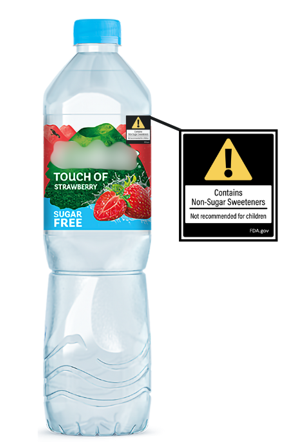 |
| 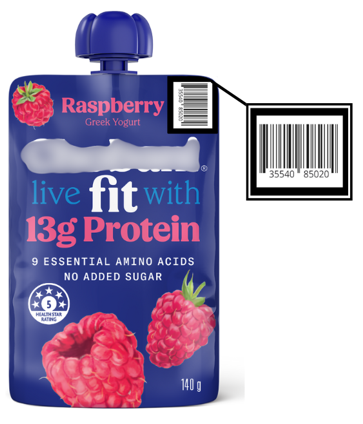 | 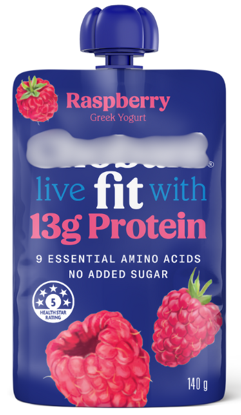 | 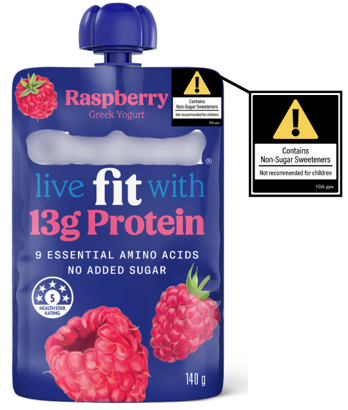 |
| 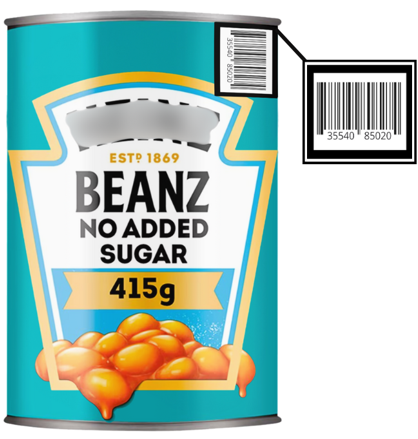 | 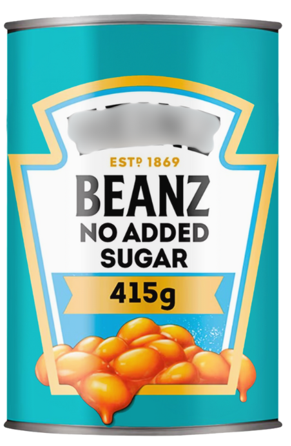 | 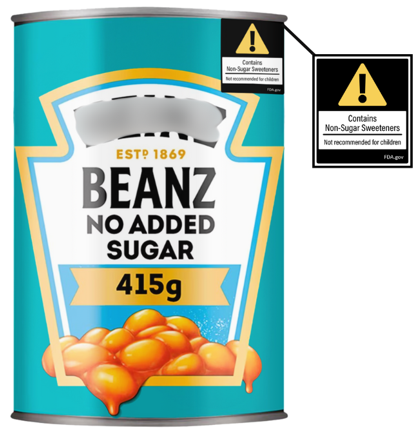 |
| Contain added sugar and NSS | | |

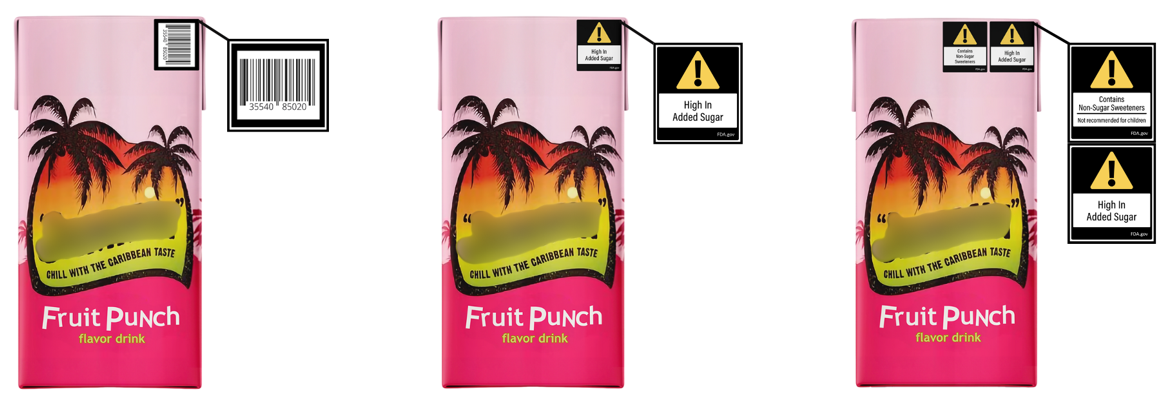

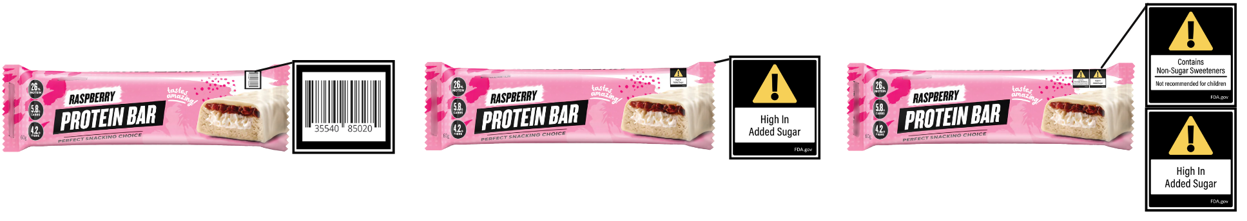

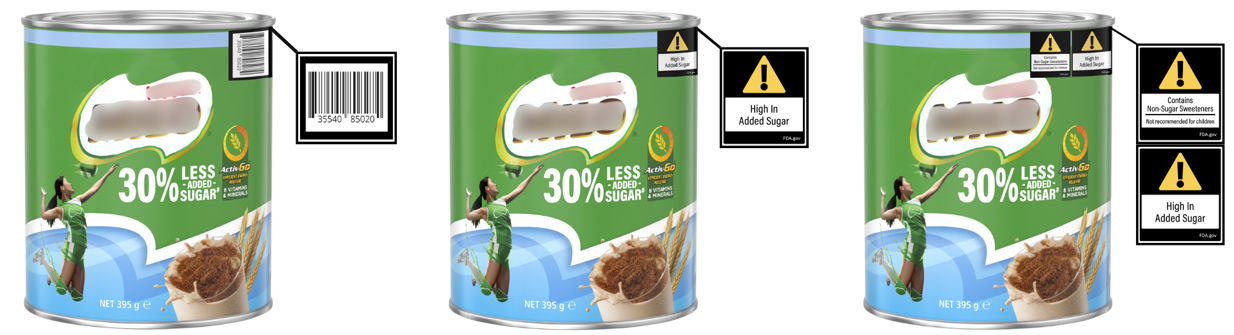
