## Supplementary figures and images for "Effects of front-of-package labels for added sugars and non-sugar sweeteners (NSS) on parents’ perceptions and selections of foods and beverages for their children: a randomized experiment"

### Supplemental Figure 4

**Supplemental Figure 4**. Participant flowchart

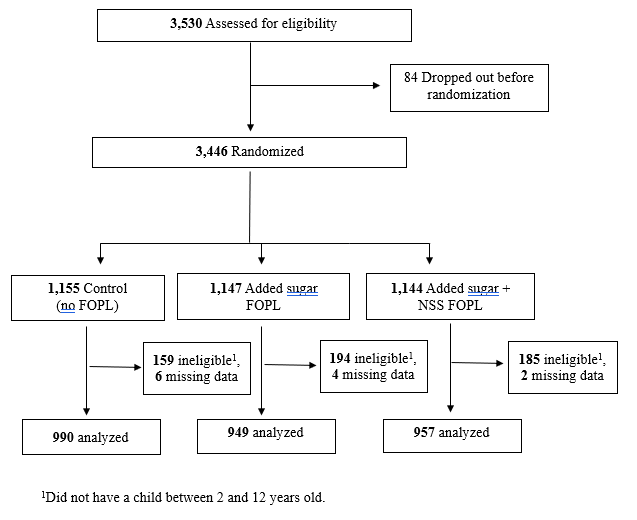
